# Conditional willingness to care for patients with Ebola virus disease: a qualitative study of risk-benefit perceptions and support needs of clinical students at a Ugandan medical school

**DOI:** 10.64898/2026.08.29.26361701

**Authors:** Bevindah Carol Nakabuubi, Racheal Nabunya, Tom Denis Ngabirano, Joyce Nankumbi, Lydia Kabiri, Edwin Kigozi, Auma Christine, Daniel Musindi, Isaac Alinda, Ammon Mark Kyokwijuka, Patience Muwanguzi

## Abstract

**Introduction:** Clinical students are a future health workforce, yet their roles during outbreaks of highly infectious diseases remain uncertain because of safety, training, supervision and welfare concerns. Uganda’s 2022 outbreak of Ebola disease caused by Sudan ebolavirus highlighted the need to understand how clinical students perceive outbreak-related care.

**Aim:** This study explored willingness to care for patients with Ebola virus disease among clinical students at a Ugandan medical school and examined how perceived risks, perceived benefits and support needs shaped that willingness.

**Methods:** An exploratory descriptive qualitative study was conducted among clinical students of Makerere University in Kampala, Uganda. Fifteen undergraduate medical and nursing students in the later years of training were purposively selected. Data were collected through in-depth interviews, audio-recorded with consent, transcribed verbatim, de-identified and analysed using latent content analysis. The Health Belief Model sensitised interpretation, and reporting was strengthened using the COREQ guidance.

**Results:** Five interrelated themes emerged, showing that willingness to care was conditional rather than simply present or absent. Students described an initial willingness grounded in professional duty, devotion to patients and the desire to save life. This willingness was restrained by perceived risks of contracting Ebola virus disease, dying, transmitting infection to family members or colleagues, emotional distress, lack of epidemic-readiness in the curriculum, inadequate preparedness and weak welfare support. Perceived benefits, including patient survival, professional learning, outbreak experience and personal fulfilment, strengthened willingness but did not override safety concerns. Students identified reliable personal protective equipment, epidemic-ready curricula, practical infection-prevention and control training, simulation, clear protocols, close supervision, psychosocial support, insurance and fair compensation as cues to action that could convert willingness into safe participation.

**Conclusions:** Clinical students in this Ugandan teaching hospital expressed a strong sense of professional responsibility, but their willingness to participate in Ebola care was conditional upon preparedness, protection, epidemic-ready education and institutional trust. Professional duty and learning opportunities promoted participation, whereas perceived risks and inadequate support limited it. Medical education programmes and outbreak-response systems should develop ethical, supervised, competency-based student roles supported by practical curricula, reliable protective equipment and psychosocial and welfare safeguards.

## Background

Ebola virus disease (EVD) is a severe infectious disease associated with substantial mortality, fear, stigma and disruption to health services [1, 2]. During the 2022-2023 Uganda outbreak caused by Sudan ebolavirus, Uganda reported 164 cases, including 142 confirmed and 22 probable cases, 55 confirmed deaths and 87 recoveries [3]. At least 19 health workers were infected and seven died [4]. These figures illustrate the occupational risks faced by health personnel during Ebola outbreaks and the importance of preparing the health workforce pipeline before the next emergency.

Health professional students occupy a complex position as a potential reserve workforce during infectious-disease emergencies, when health systems may face staff shortages, increased workloads and disruption to routine services. They are learners rather than fully licensed practitioners, yet they are embedded in clinical settings and represent future workforce capacity. Teaching hospitals and universities must therefore balance patient care needs, student safety, continuity of education, professional formation and ethical obligations to patients. During the COVID-19 pandemic and other infectious-disease emergencies, studies of medical and nursing students found that willingness to work or provide care was shaped by knowledge, clinical experience, perceived preparedness, moral responsibility, fear of infection, family safety and the availability of personal protective equipment (PPE) [5–8].

Evidence on willingness to provide care during outbreaks has grown, but there remains limited qualitative evidence from African teaching hospitals and from diseases such as Ebola, where biosafety requirements, case fatality, fear and stigma are especially salient. Evidence from Ebola-related knowledge, attitudes and preparedness studies suggests that willingness to provide care is closely linked to perceived personal safety, access to PPE, practical IPC training, confidence in institutional preparedness and credible risk communication [2, 9].

Rather than treating willingness as a simple yes-or-no position, this study understood willingness as conditional and shaped by how clinical students weighed professional duty and perceived benefits against perceived susceptibility, disease severity, barriers to care, and cues to action such as training, supervision and availability of PPE [10, 11]. Understanding these judgements is important for medical education because future health workers may be expected to contribute to emergency response, yet poorly prepared involvement could expose students, patients and communities to harm.

This conditional framing is also important because duty-to-care is rarely experienced in isolation from students’ social responsibilities. Studies of nurses and health workers involved in or preparing for Ebola response show that willingness may be moderated by fear of transmitting infection to family members, anticipated stigma, psychological distress and trust that institutions will provide clear protocols, reliable equipment and fair support [12, 13].

These concerns are directly relevant to clinical education, where clinical students require both ethical formation and practical safeguards before participating in high-risk outbreak care. The absence of clear policies defining student roles during high-risk outbreaks creates ethical and operational uncertainty for educational institutions and health systems. Understanding students’ perspectives is therefore essential for developing outbreak-preparedness strategies that balance educational needs, workforce demands and learner safety.

The aim of this study was to explore willingness to care for patients with Ebola virus disease among clinical students at a Ugandan medical school, and to describe how perceived risks, perceived benefits and institutional support needs shaped that willingness.

## Methods

### Study design and reporting

This was an exploratory descriptive qualitative study. The design was selected because the study sought to generate an in-depth account of clinical students’ perceptions, meanings and support needs in relation to a context-specific issue where limited empirical evidence was available. In-depth interviews were used to explore willingness to care for patients with EVD, perceived risks and benefits of providing such care, and practical measures that could increase students’ willingness in future outbreaks. The manuscript was prepared with reference to the Consolidated Criteria for Reporting Qualitative Research (COREQ), a 32-item checklist for interviews and focus groups [14].

### Theoretical orientation

An interpretivist qualitative orientation underpinned the study, recognising that willingness is shaped by participants’ meanings and the institutional context in which those meanings are formed. The Health Belief Model provided a sensitising framework for interpretation [15].

The model was appropriate because it focuses on how action is influenced by perceived susceptibility, perceived severity, perceived benefits, perceived barriers, cues to action and modifying factors. In this study, the model helped organise how clinical students weighed the threat of Ebola, the perceived value of patient care and professional learning, and the practical conditions needed to participate safely. Coding remained inductive, while the model informed the higher-level interpretation of how risk and benefit perceptions shaped willingness.

### Research team and reflexivity

The original study was conducted by BCN as the principal investigator for a dissertation, under the supervision of PM, who has experience in both caring for patients with Ebola virus disease and qualitative research. The principal investigator conducted the interviews and led initial data handling and analysis. She was a nursing student and therefore shared professional experiences with participants. This facilitated rapport and understanding of clinical training environments but may also have influenced how participants framed their responses.

Reflexive discussions were held with the supervisor throughout analysis to challenge assumptions and strengthen interpretive rigour. Faculty members with qualitative research experience (RN and LK) contributed to coding discussions, categorisation and theme development to strengthen credibility. Participants were informed about the study purpose and the voluntary nature of participation before consent was obtained.

### Study setting and participants

The study was conducted at Makerere University College of Health Sciences in Kampala, among students in clinical years. The study population comprised clinical medical and nursing students in the later years of training. Eligible participants were students who consented to participate, were undertaking clinical rotations at the study site, and were in the third, fourth or fifth year of training. Students who were not available in the study area during data collection were excluded.

Purposive sampling was used to select participants who could provide rich information relevant to the research questions [16]. Maximum variation was sought across programme and year of study [17]. Recruitment continued until data saturation was reached, meaning that no substantially new information was emerging from subsequent interviews [18]. The final sample included 15 participants.

### Data collection

Data were collected using face-to-face in-depth interviews conducted in a private setting. Before each interview, the researcher explained the study purpose and procedures, addressed participants’ questions and obtained written informed consent. Participants were also asked for permission to audio-record the interview. The interview guide included open-ended questions about perceived risks of caring for patients with EVD, perceived benefits of providing care, willingness to participate in care during a future outbreak, factors influencing willingness or reluctance, and support or resources that would increase willingness. Example questions included: “*What specific risks do you perceive when caring for patients with Ebola virus disease?*” and *“What support or resources would increase your willingness to provide care to patients with Ebola virus disease?”*

The interview guide was pre-tested with clinical students at the study site and adjusted to improve clarity. Interviews lasted approximately 40-60 minutes. After each interview, the researcher addressed any concerns raised by the participant and thanked the student for participating. Repeat interviews were not reported in the source study documentation.

### Data management and analysis

Audio-recorded interviews were transcribed verbatim and de-identified before analysis. Transcripts and recordings were stored securely on a password-protected computer and were accessible only to the research team. Data were analysed using latent content analysis, which enabled interpretation of both explicit content and underlying meanings in participants’ accounts [19]. The analysis involved repeated reading of transcripts, writing analytical notes and reflections, identifying meaning units, coding, comparing codes with the original text, grouping related codes into categories and developing themes. This was manual coding.

The initial coding tree included willingness, unwillingness, risk perception and benefit perception. For this manuscript, the themes were refined to improve clarity and to reflect willingness as the central organising concept. Risk and benefit perceptions were therefore presented as explanatory themes that shaped willingness rather than as separate, parallel outcomes. The refined analysis identified five interrelated themes: professional duty created an initial willingness to care; perceived risk restrained willingness through fear of infection, death and onward transmission; perceived benefits strengthened willingness through patient survival, learning and professional growth; gaps in epidemic-ready education limited confidence and willingness; and preparedness and institutional support were decisive cues to action.

### Trustworthiness

Trustworthiness was supported through several strategies [20]. Data were collected in a private setting to encourage open discussion. The interview guide was pre-tested and refined.

Interviews were audio-recorded with consent and transcribed verbatim. Transcripts were checked for accuracy and de-identified. Codes and categories were compared with the original text to preserve meaning. Theme development involved discussion with faculty members experienced in qualitative research. Direct quotations with participant identifiers are presented in the results to show the relationship between the data and interpretations.

## Results

### Participant characteristics

Fifteen students participated in the study. Seven participants (47%) were male. Eight participants (53%) were undergraduate nursing students, while seven (47%) were medical students. Three participants (20%) were in third year, nine (60%) were in fourth year, and three (20%) were in fifth year.

### Conditional willingness to care for patients with EVD

Five themes emerged regarding the conditional willingness of clinical students to care for patients with EVD, as shown in the results framework in Figure 1 and expanded in the results section. The framework shows that willingness was shaped by professional duty, perceived benefits, perceived risks, epidemic-ready education and institutional support. Practical training, reliable PPE, supervision, clear protocols and welfare support could strengthen willingness and enable safer student participation in outbreak care.

**Figure 1:**
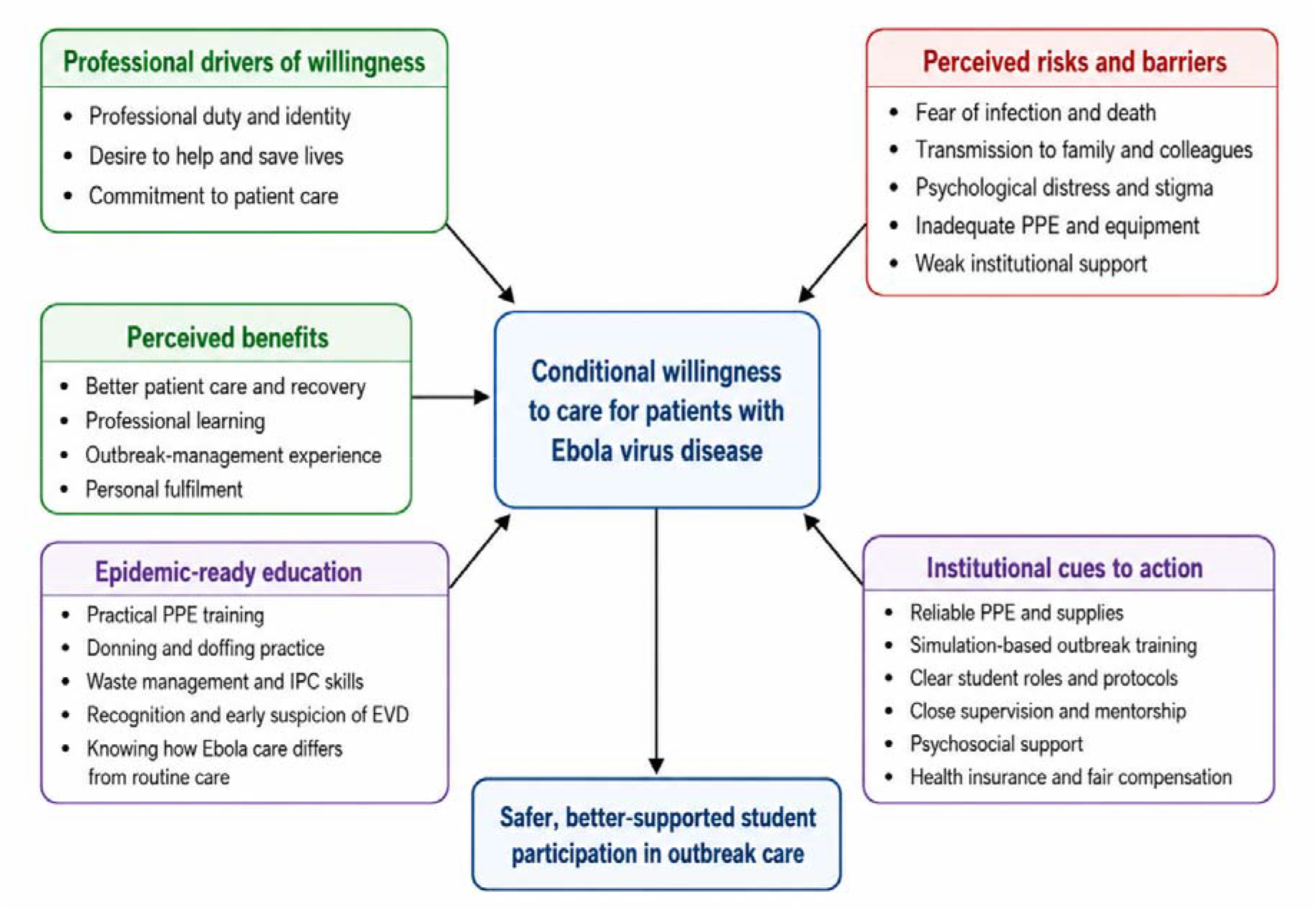
Results framework: Conditional willingness of clinical students to care for patients with EVD.

### Theme 1: Professional duty created an initial willingness to care

Many participants described an initial willingness to care for patients with EVD. This willingness was grounded in professional identity, moral responsibility and devotion to helping patients. Students described health professional training as a commitment to serve, even when care involved personal risk. Participants expressed this duty through statements that framed care as a calling, prioritised saving life and rejected allowing fear to prevent life-saving action:

*“…medicine is a calling….”* (Participant 10, medicine student)

*“….saving life is a priority….”* (Participant 1, Medicine student)

*“I would not want to lose a life because I am scared.”* (Participant 8, nursing student)

However, this willingness was not absolute. Students did not present willingness as heroic self-sacrifice detached from safety. Instead, they framed it as a professional responsibility that should occur within a prepared and protective system. Thus, willingness began with professional duty but was negotiated against practical questions about training, PPE, supervision and institutional support.

### Theme 2: Perceived risk restrained willingness through fear of infection, death and onward transmission

All participants recognised EVD care as risky. The main perceived risk was contracting infection through direct contact with a patient, body fluids or a contaminated clinical environment. Students understood Ebola as highly infectious and potentially fatal, and some linked this fear to awareness that health workers had been infected or had died during outbreaks. For students who had not received practical simulation or direct experience of Ebola care, fear was intensified by uncertainty about what to do and concern about making mistakes.

One participant described the basic occupational risk in direct terms:

> *“Of course, there are risks because this is an infectious disease. In case you come in contact with an infectious person you can get infected, and you can transmit the disease to infect other people.”* (Participant 13, nursing student)

Risk was also relational. Students worried not only about their own safety but also about becoming a source of infection for colleagues, other patients, household members and communities. The possibility of carrying infection home was a major concern and reduced willingness to participate unless strong infection-prevention measures were in place.

> *“…but then risk of transmitting it from this person that has come with it and then me getting it and taking it to my colleagues or even where I could be staying…”* (Participant 4, medicine student)

Perceived risk also included psychological and emotional consequences. Students referred to fear, anxiety, guilt, stigma, trauma and possible post-traumatic stress. This suggests that EVD preparedness for students should include psychosocial readiness and support, not only technical infection-prevention training.

### Theme 3: Perceived benefits strengthened willingness through patient survival, learning and professional growth

Although participants perceived EVD care as risky, they also identified important benefits. Patient-related benefits included symptom management, recovery, survival and return to family. Students felt that patients with EVD should not be abandoned because of fear and that good care could give patients hope and another chance to live. One participant emphasised the patient’s potential gain from care:

> *“…On the patient side, the patient has everything to gain because they can go back to their families and live their lives…so they have all to gain because it is a deadly disease and if you survive there’s a story to tell at the end of the day.”* (Participant 15, nursing student)

Professional benefits were also important. Students saw outbreak participation as a rare learning opportunity that could improve their competence in infection prevention, emergency response and management of highly infectious diseases. The perceived educational value of caring for patients with EVD strengthened willingness, particularly when students imagined being supervised by experienced staff.

> *“I believe one direct benefit would [be] we’ll be learning the management of such a case, which is helpful be it in the outbreaks whether it’s Ebola or any other infectious disease generally.”* (Participant 7, nursing student)

Personal benefits included fulfilment, confidence and satisfaction from contributing to recovery. However, participants did not suggest that benefits alone were enough to justify exposure. Rather, perceived benefits strengthened willingness only when combined with adequate preparedness and protection.

### Theme 4: Gaps in epidemic-ready education limited confidence and willingness

A further barrier to willingness was the perceived gap between classroom knowledge about EVD and practical readiness to work in an Ebola treatment unit. Participants felt that they had been taught about the disease but not sufficiently prepared for the practical realities of outbreak care, including donning and doffing PPE, waste management, triage, escalation, and how care in an Ebola treatment unit differs from routine clinical care. This lack of practical epidemic-readiness increased anxiety because students felt they might not know what to do if called upon to participate.

One participant linked their reluctance directly to this curricular gap:

> *“One of my biggest barriers for working with Ebola patients is that I don’t know what to do. We’ve been taught about the disease in class, but we didn’t learn how to don and doff PPE…what you do when you get there…Basically, our curriculum needs to be epidemic-ready.”* (Participant 2, medicine student)

Participants suggested that epidemic-readiness should be built into health professional curricula before outbreaks occur, rather than being introduced only once a disease has already emerged. They wanted practical sessions that would help them recognise suspected cases, maintain a high index of suspicion, use PPE correctly, manage clinical waste safely and approach outbreak care with confidence rather than panic.

### Theme 5: Preparedness and institutional support were decisive cues to action

Students repeatedly described willingness as dependent on the conditions under which they would be expected to care for patients. Reliable PPE, practical training, clear protocols, mentorship, close supervision, medical equipment, compensation and insurance were seen as cues to action that could convert willingness into safe participation.

PPE was central. One participant explicitly linked willingness to the availability of protective equipment and the ability to use clinical skills safely:

> *“One of the factors would be if there are provided resources, that’s the protective equipment, like gloves, masks, gowns, gumboots, to ensure my safety, then I would be willing to participate because I would use the skills, and the knowledge to care for the patient.”* (Participant 11, nursing student)

In contrast, shortages of PPE and medical equipment, improvisation and weak institutional support reduced willingness and created emotional distress. One participant linked inadequate supplies to compromised care and psychological strain:

> *“Thirdly, when you’re trying to provide care to the Ebola patient you may find that you don’t have enough protective equipment, medical equipment to offer the best patient care…There was a lot of improvising, and patients were asked to buy a cannula, syringes, and gloves which affected the kind of care we offered to those patients which could tapper with you psychologically.”* (Participant 3, medicine student)

Participants also identified the need for simulation-based training, especially in donning and doffing PPE, infection prevention and control, triage, supportive care and escalation pathways. They wanted close supervision by experienced clinicians and clear policies defining whether, when and how students could safely participate. Welfare concerns also mattered. Some students referred to previous experiences in which health workers had reportedly not received promised allowances or compensation, creating mistrust. Insurance, compensation and transparent support arrangements were therefore not simply incentives.

They were perceived as evidence that institutions valued and protected those taking risks.

## Discussion

This study explored willingness to care for patients with Ebola virus disease among clinical students at a Ugandan medical school. The main finding is that willingness was conditional. Students expressed a strong sense of professional duty and recognised benefits for patients and their own professional development, but their willingness depended on perceived safety, preparedness, supervision, PPE and institutional trust. This framing is stronger than presenting willingness, risk perception and benefit perception as separate findings because the data show that risk and benefit perceptions operated through willingness: they either strengthened, restrained or qualified students’ readiness to participate.

The finding that willingness was shaped by preparedness and PPE is consistent with evidence from health professional students and health workers during infectious-disease outbreaks. Patel and colleagues found that health care students’ willingness to work during infectious-disease outbreaks was associated with knowledge, disaster training and PPE availability [5]. Evidence from Ebola-related KAP and preparedness studies in Ethiopia, Ghana and Nigeria similarly links knowledge, IPC training, PPE access and perceived readiness with more favourable attitudes toward Ebola response [12, 21, 22]. A recent nationwide survey among emergency healthcare workers in Uganda also found varied readiness to manage EVD and highlighted demand for further training in EVD management, IPC, clinical presentation, vaccination and diagnosis [23]. In this Ugandan student study, PPE was not a minor logistical issue. It represented the boundary between professional duty and unacceptable risk.

A distinct contribution of this study is that students located preparedness not only in the hospital or Ebola treatment unit, but also in the curriculum. Participants felt that classroom teaching on Ebola diagnosis and management had not been matched by practical preparation for outbreak work, including donning and doffing PPE, waste management, triage, escalation and care in an Ebola treatment unit. This supports the need for medical and nursing curricula to include repeated, competency-based epidemic-readiness training before outbreaks occur. Evidence that structured educational interventions can improve Ebola-related knowledge and attitudes among frontline workers [9, 24], and that emergency health workers in Uganda have reported a need for further EVD training [25, 26], reinforces the importance of curricula that link disease knowledge to practical action.

Professional identity was a key motivator. Participants described health professions as involving a duty to save life and respond to patients in need. This aligns with literature showing that students may express moral responsibility to care even when they are anxious or uncertain [6, 7]. However, the findings caution against relying on duty alone. Studies among nurses after admission of a patient with EVD show that willingness to provide care may involve moral tension between professional duty and obligations to protect family members [27]. Students’ willingness was strongest when professional responsibility was paired with institutional protection. Medical education should therefore avoid narratives that romanticise sacrifice and should instead emphasise ethical service under safe, supervised and adequately resourced conditions.

Fear of infection and onward transmission to family members was central to perceived risk. This is important because students did not perceive risk as purely individual. They feared becoming a route of transmission to colleagues, patients, household members and communities. Similar concerns have been documented among health workers and students during Ebola, influenza and COVID-19 outbreaks [8, 28, 29]. The broader epidemic-risk literature also shows that risk perception is shaped by trust, information sources, perceived control and the credibility of authorities [30]. Outbreak-preparedness training for clinical students should therefore include not only PPE skills but also family-safety counselling, decontamination routines, risk communication and strategies for reducing stigma [31].

Participants also identified psychological distress as a barrier to willingness. The possibility of guilt, stigma, trauma and post-traumatic stress suggests that student preparedness should include emotional readiness and access to psychosocial support. Experiences from Ebola responses in West Africa have shown that health workers can face substantial emotional burden, fear, stigma and social consequences [32, 33]. Work on the mental-health impact of the 2014-2016 West African Ebola epidemic further shows that fear and culture shaped psychological and social responses to the outbreak [13]. Teaching hospitals should therefore integrate debriefing, peer support and referral pathways into outbreak-related student policies.

Beyond preparedness and institutional support, the findings raise ethical and policy questions about the role of clinical students during infectious-disease outbreaks. Participants’ accounts highlighted a tension between educational participation and learner safety. Student involvement should therefore remain voluntary, be limited to tasks for which competence has been demonstrated, and occur only with adequate training, supervision, protective equipment and clear escalation pathways. Institutions should also define alternative learning arrangements for students who do not participate in direct high-risk care.

The findings have practical implications for medical education in Uganda and similar settings. First, epidemic-readiness should be embedded in the curriculum before emergencies occur, including repeated practical simulation in donning and doffing PPE, infection prevention and control, waste management, triage, supportive care and escalation. Training interventions, including mobile and targeted educational approaches, have improved Ebola-related knowledge and attitudes among frontline workers, suggesting that structured preparation can strengthen confidence as well as competence [22]. Second, institutions should define ethical student roles during high-risk outbreaks, including criteria for exclusion from direct care, supervised participation, alternative learning arrangements and academic continuity. Third, students’ willingness may be strengthened by visible institutional commitments to PPE, supervision, psychosocial support, insurance, fair compensation and transparent risk communication [12, 23, 27, 30]. Finally, students should be involved in preparedness planning, because their concerns identify the conditions under which future workforce members can contribute safely and meaningfully.

### Strengths and limitations

A strength of this study is that it provides context-specific qualitative evidence from clinical students in a Ugandan teaching hospital following a national Ebola outbreak. The use of indepth interviews allowed students to describe nuanced perceptions of risk, benefit, duty and institutional support. The revised analysis and reporting were strengthened using COREQ, and the results include direct quotations with participant identifiers to support transparency.

The study also has limitations. It was conducted at one teaching hospital and involved a small qualitative sample, which limits transferability. The final sample included medical and nursing students; views from other clinical programmes may not have been fully represented. Willingness was explored as a stated intention rather than observed behaviour during an active outbreak, and students’ actions during a real emergency may differ from interview accounts. Some COREQ items were not fully documented in the available source materials, including interviewer gender, prior relationship with participants, refusal rates, whether non-participants were present, whether transcripts were returned and whether member checking was conducted. These should be verified by the study team before submission.

### Conclusions

Clinical students at this Ugandan medical school expressed willingness to care for patients with Ebola virus disease, but this willingness was conditional. Professional duty, devotion to patients, patient survival, learning and professional growth strengthened willingness. Fear of infection, death, onward transmission, emotional distress, gaps in epidemic-ready education, inadequate preparedness and weak welfare support restrained it. Practical cues to action, including reliable PPE, epidemic-ready curricula, simulation-based training, clear protocols, close supervision, psychosocial support, insurance and fair compensation, were central to converting willingness into safe participation. Medical education programmes and teaching hospitals should establish competency-based participation frameworks and provide adequate institutional support to enable meaningful involvement while safeguarding learners and the communities they serve.

## Declarations

### Ethics approval and consent to participate

Ethical approval was obtained from the Makerere University School of Health Sciences Research and Ethics Committee (ref number MakSHSREC-2023-502). Administrative clearance was obtained from Mulago Hospital (ref number MHREC 2543). Written informed consent was obtained from all participants before data collection. Participation was voluntary, and participants were informed that they could withdraw at any time without disadvantage. Data were de-identified to protect confidentiality. The study was governed by the Helsinki declaration principles.

### Consent for publication

The manuscript contains de-identified participant quotations and does not include identifiable individual participant data. All co-authors consented to the publication.

### Availability of data and materials

The qualitative datasets generated and analysed during the current study are not publicly available because they contain information that could compromise participant confidentiality. De-identified data may be available from the corresponding author on reasonable request and subject to relevant ethical approvals.

### Authors’ contributions

BCN conceptualised the study, collected data, participated in analysis and prepared the original dissertation report. PAM supervised the study and contributed to study design, interpretation and manuscript development. RN, TDN, LK, EK and AC contributed to manuscript development and critical review. All authors reviewed and approved the final manuscript before submission.

## Data Availability

The qualitative datasets generated and analysed during the current study are not publicly available because they contain information that could compromise participant confidentiality. De-identified data may be available from the corresponding author upon reasonable request and subject to relevant ethical approvals.

## Acknowledgements

The authors acknowledge the Department of Nursing, College of Health Sciences, Makerere University; Mulago National Referral Hospital; and the clinical students who participated in the study. Permission should be obtained from any individuals named in the acknowledgements before submission.

## List of abbreviations

BDS: Bachelor of Dental Surgery
BNUR: Bachelor of Nursing
COREQ: Consolidated Criteria for Reporting Qualitative Research
COVID-19: Coronavirus disease 2019
EVD: Ebola virus disease
HBM: Health Belief Model
IPC: Infection prevention and control
MakSHSREC: Makerere University School of Health Sciences Research and Ethics Committee
MBChB: Bachelor of Medicine and Bachelor of Surgery
PPE: Personal protective equipment
SUDV: Sudan ebolavirus
WHO: World Health Organization

